# A machine learning approach to identifying important features for achieving step thresholds in individuals with chronic stroke

**DOI:** 10.1101/2022.01.27.22269966

**Authors:** Allison E. Miller, Emily Russell, Darcy S. Reisman, Hyosub E. Kim, Vu Dinh

**Author notes:** Corresponding author (AM). These authors contributed equally to this work (AM and ER are co first authors). These authors contributed equally to this work (HEK and VD are co senior authors).

## Abstract

**Background:** While many factors are associated with stepping activity after stroke, there is significant variability across studies. One potential reason for this variability is that there are some characteristics necessary to achieve greater stepping activity that differ from others that may need to be targeted to improve stepping activity.

**Objective:** Using two step thresholds (2500 steps/day, corresponding to home vs. community ambulation and 5500 steps/day, corresponding to achieving physical activity guidelines through walking), we applied 3 different algorithms to determine which predictors are most important to achieve these thresholds.

**Methods:** We analyzed data from 268 participants with stroke that included 25 demographic, performance-based and self-report variables. Step 1 of our analysis involved dimensionality reduction using lasso regularization. Step 2 applied drop column feature importance to compute the mean importance of each variable. We then assessed which predictors were important to all 3 mathematically unique algorithms.

**Results:** The number of relevant predictors was reduced from 25 to 7 for home vs. community and from 25 to 16 for aerobic thresholds. Drop column feature importance revealed that 6 Minute Walk Test and speed modulation were the only variables found to be important to all 3 algorithms (*primary characteristics)* for each respective threshold. Other variables related to readiness to change activity behavior and physical health, among others, were found to be important to one or two algorithms (*ancillary characteristics)*.

**Conclusions:** Addressing physical capacity is *necessary but not sufficient* to achieve important step thresholds, as *ancillary characteristics,* such as readiness to change activity behavior and physical health may also need to be targeted. This delineation may explain heterogeneity across studies examining predictors of stepping activity in stroke.

## Introduction

Stroke is a leading cause of disability world-wide and results in numerous sequelae, including reduced walking ability and aerobic deconditioning [1, 2]. This is problematic because reduced walking ability and aerobic deconditioning are associated with deficits in physical function [3, 4], depression [5, 6], and reduced self-efficacy [7].

As a result, many individuals with stroke are inactive [8] and not meeting physical activity recommendations to maximize health benefits [9, 10]. In parallel, individuals with stroke often report improving their walking ability as a primary goal for rehabilitation [11] and clinicians spend considerable time on interventions to improve their walking [12].

Thus, two areas of particular relevance for the rehabilitation community are determining predictors of daily stepping activity that may inform whether an individual with stroke will be able to walk in the community or if they will be primarily home bound [3, 13, 14] and whether they will meet aerobic activity guidelines through walking [10, 15]. The latter is salient for clinicians as reaching physical activity recommendations may have implications for future health outcomes [16–19].

Previous work has suggested that ∼2500 daily steps distinguishes between home versus community ambulators in individuals with stroke [3] and that ∼5500 daily steps is a reasonable target for individuals with disabilities to meet physical activity guidelines [10]. However, there has been significant heterogeneity in predictors of daily stepping activity after stroke. A recent meta-analysis including 26 studies and over 30 predictors of stepping activity post stroke found inconsistencies in the relevance of certain predictors [20]. This finding, in conjunction with the limited efficacy of interventions targeting daily stepping activity post stroke [21], suggests a need to better understand which predictors are most important for improving daily stepping activity after stroke.

To this end, the large number of variables that may influence walking activity after stroke requires analytical techniques with the ability to handle large, heterogeneous datasets. Machine learning techniques have this capacity as well as other advantages, including requiring fewer assumptions about the distributions of the data, numerous options for non-parametric models and dimensionality reduction techniques, and most notably their strong predictive capabilities [22–24]. Recent work has utilized machine learning to predict recovery of upper limb functioning [23, 24] and functional outcomes after stroke with high accuracy [25]. In particular, one approach to determining which predictors may be most relevant is to utilize multiple different machine learning algorithms and compare predictors across algorithms [24]. This approach can help gain a better understanding of a target population and increase predictive power, as features that are shown to be important across mathematically unique algorithms likely represent fundamental characteristics of that population and are therefore important in predicting the outcome. Said differently, comparing relevant predictors across mathematically unique algorithms may help differentiate predictors that are *most important* for predicting the outcome from predictors that *might* be important for predicting the outcome in some individuals.

Despite these clear advantages, principled application of ML algorithms has not been used to understand the most important characteristics of stroke survivors who achieve stepping thresholds for community mobility status [3] or physical activity recommendations [10]. Thus, we had two objectives for this study. First, we aimed to apply three mathematically unique algorithms to a large dataset to predict achievement of community ambulation status (2500 steps/day) [3] and/or meeting physical activity recommendations (5500 steps/day) [10] and compare predictors across algorithms. We were specifically interested in identifying predictors that were found to be important to all three algorithms. Based on previous studies, we hypothesized that measures of physical capacity, specifically gait endurance [3, 26] (6 Minute Walk Test) and gait speed [14, 27] (10-Meter Walk Test), would be important predictors to all three algorithms and that balance self-efficacy [28, 29] (Activities Specific Balance Confidence Scale) and environmental factors [30–33] (Area Deprivation Index), would be important predictors to one or two algorithms but not all three. Our second objective was to assess the prediction accuracy of these three different machine learning algorithms for each threshold.

## Methods

### Participants

Data was obtained from the baseline timepoint of a randomized clinical trial comparing the efficacy of specific interventions for improving daily walking activity in individuals with stroke [34]. Table 1 lists the eligibility criteria for this study. All participants signed informed consent approved by the Human Subjects Review Board at the University of Delaware prior to study participation (IRB #878153-50). This work has been conducted according to the principles expressed in the *Declaration of Helsinki*.

**Table 1.** Eligibility criteria

| Inclusion Criteria | Exclusion Criteria |
| --- | --- |
| Ages 21 – 85 | Cerebellar stroke |
| ≥6 months post stroke | Other neurologic conditions in addition to stroke |
| Able to walk without assistance at a gait speed of ≥0.3 m/s | Lower extremity Botulinum toxin injection < 4 months earlier |
|  | Current participation in physical therapy |
|  | Inability to walk outside the home before their stroke |
|  | Coronary artery bypass graft, stent placement, or myocardial infarction within the past 3 months |
|  | Musculoskeletal pain limiting activity |
|  | Inability to communicate with the investigators |
|  | Inability to answer at least one orientation question correctly (item 1b of the National Institutes of Health Stroke Scale) and inability to follow at least one, two-step command (item 1c of the National Institutes of Health Stroke Scale) |

### Measures

The following continuous measures were included in the statistical analysis: 6-Minute Walk Test (6MWT) [35, 36], self-selected walking speed (SSWS) [36], fastest walking speed (FWS) [36], speed modulation (calculated as FWS – SSWS), Montreal Cognitive Assessment (MoCA) [37], Charlson Comorbidity Index (CCI, age-adjusted) [38], Patient Health Questionnaire-9 (PHQ-9) [39], Activities Specific Balance Confidence Scale (ABC) [40], body mass index (BMI), age, time since initial stroke, number of strokes, number of medications (including supplements), Area Deprivation Index state decile (ADI_S) [41, 42], and Area Deprivation Index national percentile (ADI_N) [41, 42].

The following categorical measures were included in the analysis: usual orthotic device [0= no orthotic device, 1=orthotic device], usual assistive device [0= no assistive device, 1= assistive device], living situation [0= living alone, 1= living with a family member/significant other, 2= living alone but has outside assistance daily, 3= other], marital status [0= married, 1= not married], work status [0= employed full-time, 1= employed part-time, 2= retired, 3= unemployed (includes being on disability)], years of education [0= high school (≤15 years), college graduate (16 years), above graduate (>16 years)], gender [0= male, 1= female], side of hemiparesis [0= left, 1= right, 2= bilateral], readiness to change activity behavior [1= currently not active and do not intend on becoming active in the next 6 months, 2= currently not active but thinking about starting to become active in the next 6 months, 3= currently active sometimes but not regularly, 4= currently active regularly but have only begun doing so within the last 6 months, 5= currently active regularly and have done so for longer than 6 months], and relapse in activity behavior [1= experienced a relapse in activity levels, 2= no relapse in activity levels] [43, 44].

### Daily stepping activity

To measure daily stepping activity, participants were provided with a FitBit^TM^ at the baseline visit of the clinical trial. The FitBit^TM^ has acceptable accuracy in detecting stepping activity in individuals with stroke [45–48]. Participants wore the device on their non-paretic ankle and were instructed to wear it for 7 days to reliably estimate daily stepping activity [49] and continue with their usual activity. Average steps per day (ASPD) was calculated by summing the total number of steps taken over all valid recording days and dividing this sum by the number of valid recording days.

### Statistical analysis

#### Data processing

Figure 1 displays a data pipeline that describes the data and analysis procedures. First, the data was exported from the electronic database, REDCap [50], and initially comprised 283 individuals and 32 clinical and demographic variables. ASPD was used to determine stepping thresholds (home vs. community threshold of 2500 steps/day and the aerobic threshold of 5500 steps/day). After removing variables with more than 5% missing entries and variables that were irrelevant to the outcome (e.g., “patient ID”), 25 of the remaining 32 variables were used in our analyses. Of the 283 participants, 15 were excluded for having missing data in one or more of the 25 variables selected, leaving a total of 268 participants included in our analyses. Two individuals inspected the data for accuracy prior to analysis.

**Fig 1.**
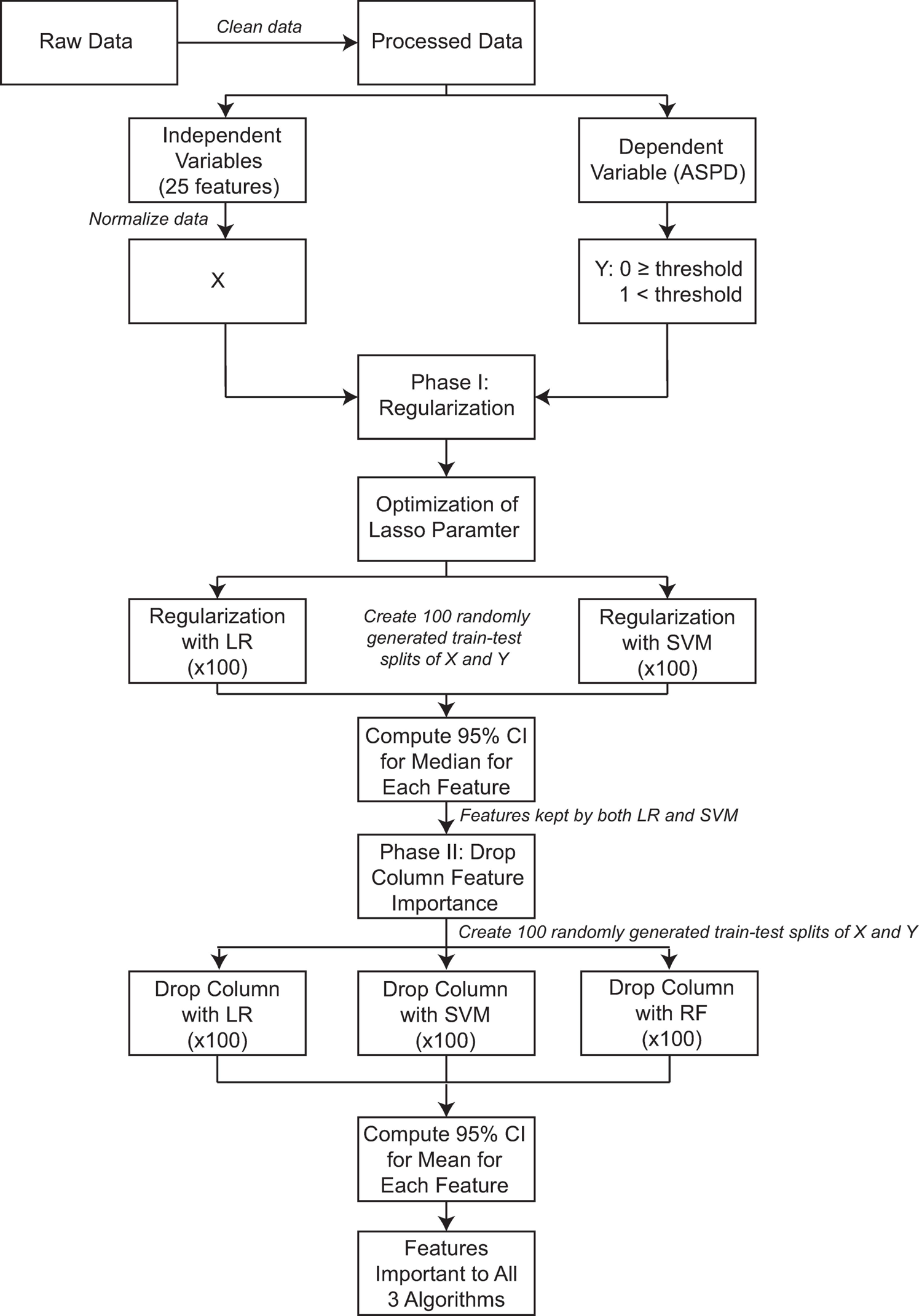
Data pipeline. Abbreviations: ASPD-Average Steps/Day, LR-Logistic Regression, SVM-Support Vector Machine, RF-Random Forest, CI-Confidence Interval

All analyses were conducted using custom-written code in the Python programming language and compiled with Spyder4, using the standard machine learning library sklearn [51]. The same procedures were repeated for both the home vs. community and aerobic thresholds. Briefly, the preprocessed data set from 268 participants was imported into a data frame. Our design matrix was composed of all 25 variables except for ASPD, which was used to compute our binary outcome variable, step threshold category. All data in our design matrix was normalized for stability using a min-max scaler, which uses the minimum value and range of the distribution to shift and scale the distribution, respectively, translating to the interval [0, 1] while preserving the shape of the original distribution.

For the home versus community threshold, participants were assigned a label of 1 for home ambulator (ASPD < 2500) or 0 for community ambulator (ASPD ≥ 2500). This resulted in a distribution of 58 (21.64%) home ambulators and 210 (78.36%) community ambulators. For the aerobic threshold, those whose ASPD were below the threshold of 5500 daily steps were given a label of 1, and those who met or exceeded the minimum aerobic threshold were labeled with a 0. The distribution in this case was 185 (69.03%) below the aerobic threshold and 83 (30.97%) above the threshold.

#### Drop-column procedure for feature importance

To address the first objective of this study, a two-stage procedure was used. The first stage was dimensionality reduction using lasso regularization. The purpose of this stage was to reduce redundancy and noise in our set of variables, thus avoiding potential bias or conflation in measures of feature importance for strongly correlated variables in the second stage. The second stage involved computing a measure of feature importance for the remaining features following dimensionality reduction (Fig 1) [52]. Throughout the analysis, the performance metrics were assessed using Monte Carlo Cross-Validation (MCCV) which in each instance consisted of 100 randomly generated train-test-splits of the data where for each split, 70% of the data was used as a training set and the remaining 30% was used as the test set.

For the first stage, we used logistic regression (LR) and linear support vector machine (SVM) both with lasso regularization with optimized regularization parameters. To optimize these parameters, a grid search was used with two refinements, each around the parameter value that was found to produce the best average model performance using MCCV. Once the optimized regularization parameters were found, the following procedure was executed for both regularized models: first, the model was fit over 100 random 70-30 train-test-splits of the data and the 100 sets of weights (i.e., coefficients, see Supplementary Material for more detail) for each variable were recorded. Then, for each variable, 1000 bootstrap samples of size 100 were generated from the sample of 100 coefficients, and an empirical 95% confidence interval for the median of the coefficients was computed. Variables for which 0 was included in the 95% confidence interval for both regularized models were then dropped, and only the remaining variables would be used in the second stage.

In the second stage, a measure of feature importance was computed for three different machine learning models: LR, SVM with a radial basis function (RBF) kernel, and random forest (RF). Our criteria for choosing these three models were that they had to be commonly-used machine learning algorithms, mathematically distinct, and previously shown to perform well with clinical and biological data (see Supplementary Material for additional details) [53]. While all three of these algorithms fit this criterion, it is important to note that, unlike parametric models like LR and SVM whose models can be written down in function form, nonparametric models like RF are known for having strong predictive power, while lacking interpretability. In this way, RF could be thought of as having insight that is more complex yet could be difficult to quantify. For this stage, we aimed to use a measure of feature importance that is both easily interpretable as well as uniformly applicable across the three algorithms. We therefore chose the drop column feature importance process.

The drop column importance measure uses a chosen metric of model performance to quantify how much a given variable contributes to a model’s performance, i.e., whether the variable helps, hurts, or has no effect on the performance of the model [54]. Once the metric for performance is chosen, the drop column feature importance can be computed using any model. The idea of this procedure is to fit the model using all variables first (i.e., the *benchmark model)* and take a measure of the model’s performance using the chosen metric (i.e., the *benchmark performance*). To measure the influence of an individual feature on this metric of model performance, that feature will then be dropped from the training set and the model is then refitted. A measure of this new model’s performance, the *dropped column performance* is then taken, and that feature’s overall *importance* is taken to be the difference between the model’s performance with and without that feature, i.e.:

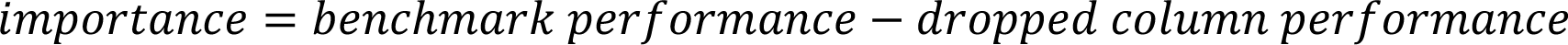

This same procedure is then followed for each feature. Thus, if a particular feature improves the model’s performance, this *importance* measure will be positive because the *dropped column performance* would be lower than the *benchmark performance* (in other words, the model performed worse when we removed that particular feature) and vice versa for features whose importance is negative. The intuition underlying this procedure is that positive features hold pertinent information about the dependent variable, as they contributed the most to correctly identifying those in the target class.

The drop column procedure was run with MCCV for all three algorithms using only the features that were retained after regularization. For consistency, a single list of 100 train-test-splits was randomly generated and used for all three algorithms in this step. For each algorithm, every feature was given a drop column importance for every train test-split, resulting in 100 importance measures associated with each algorithm for each feature. From these samples of 100 importance scores for each feature, 1000 bootstrap samples of size 100 were taken and an empirical 95% confidence interval for the mean importance score was computed for each feature. A feature was considered important to an algorithm if the 95% confidence interval for the mean importance of that feature was positive. To address our first objective, we compared predictors whose mean importance was positive across all three algorithms with the framework that predictors that met this criterion are likely critically important for predicting the outcome.

#### Model performance

For our second objective, we compared the performance of these models by examining their prediction accuracy for each step threshold. For the aerobic threshold, the metric of prediction accuracy used was standard accuracy. Due to the nature and severity of the class imbalance in the home versus community case, particularly that the target class was the minority [55], the metric of balanced accuracy, which takes the average of the recall (sensitivity) and specificity, was used to more accurately reflect the models’ performance on the target class (see Supplementary Material). With this class imbalance, balanced accuracy is better suited than standard accuracy to assess model performance because taking the average of recall and specificity takes the accurate classification of both classes into account. This avoids the case where a constant model (i.e., labeling all points as community ambulators) would result in a conflated standard accuracy score, while mis-identifying the entire target class. These models were fit using the algorithms’ default parameters from the sklearn documentation for the aerobic threshold. Due to the class imbalance in the home versus community threshold, a grid search was performed to optimize only the class weights parameter, *class_weight*, to either have the default setting of no weights or to have balanced class weights, which imposes a weight on each class during fitting that is inversely proportional to the class frequency.

To assess model performance, as well as validate the regularization step in the feature importance procedure, we computed the appropriate measure of prediction accuracy, again using MCCV over a single, newly generated set of 100 train-test-splits. This was done first using all the features, then using the set of features retained in the regularization phase. The goal of regularization is to eliminate redundant or uninformative variables, resulting in the use of fewer variables without the loss of any critical information. Achieving this would be evidenced by either no significant decrease, or even an improvement in the model’s performance when using the variables retained after regularization versus the full set of variables. The data and code associated with this work are available on Open Science Framework at https://osf.io/tgzpb/

## Results

Table 2 displays the demographic characteristics and summary step data of our full sample (n=268).

**Table 2.** Characteristics of study sample (n = 268)^a^

| Characteristic | Participant |
| --- | --- |
| Age (years) | 65 (IQR 17) |
| Gender | Male: 139 (51.9%)<br>Female: 129 (48.1%) |
| Side of Hemiparesis | Left: 142 (53%)<br>Right: 120 (44.8%)<br>Bilateral: 6 (2.2%) |
| Time Since Initial Stroke (months) | 24 (IQR 42) |
| Assistive Device (yes/no) | Yes: 126 (47%)<br>No: 142 (53%) |
| Self-selected Gait Speed (m/s) | 0.75 (IQR 0.32) |
| Average Steps per Day | 4175 (IQR 3061.5, Range 76 – 18166) |
| Total Number of Valid Stepping Days | 8 (IQR 6, Range 3 - 27) |

<sup>a</sup>Continuous variables presented as median (IQR- Interquartile range)

### Results for home versus community threshold

After the regularization process, LR and linear SVM dropped all but the same 6 features (6MWT, PHQ-9, readiness to change relapse score, usual assistive device, years of education, and ADI_N) with linear SVM also keeping 1 additional feature (SSWS). This resulted in 7 of the original 25 variables proceeding to the drop column phase.

For both LR and SVM, all 7 variables resulting from the regularization step were found to be important, with each feature contributing at least 6% improvement to the balanced accuracy score on average for both algorithms. For RF, only 1 of the 7 variables was found to be important (6MWT). Thus, the only feature that was important to all three algorithms for the home versus community threshold was 6MWT, suggesting that walking endurance is critically important for predicting community mobility status in stroke. Figure 2 displays the results of the drop column phase for this threshold.

**Fig 2.**
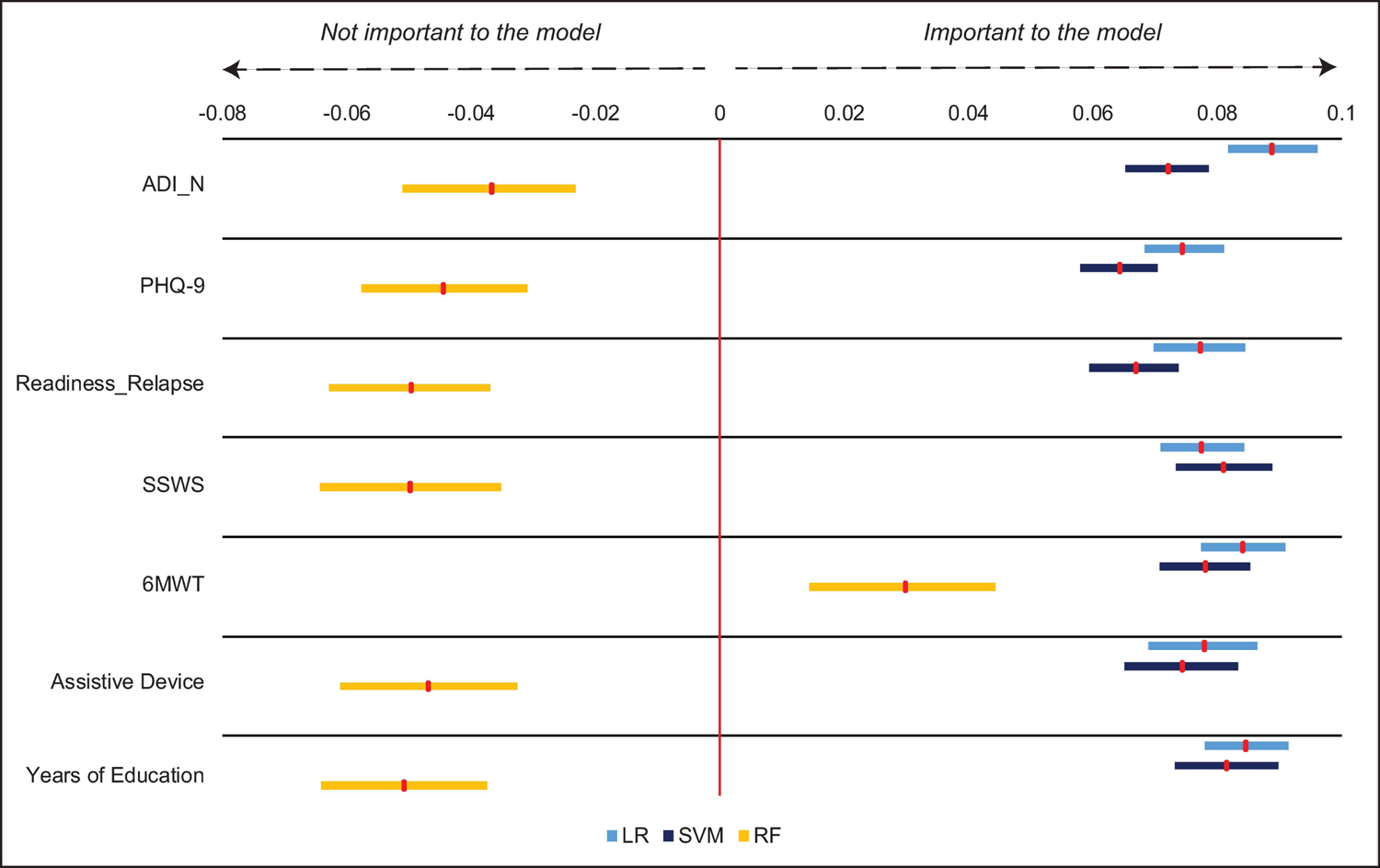
Drop column feature importance for home vs. community threshold (2500 steps/day). Red markers show mean feature importance with 95% bootstrapped confidence interval. 6MWT was the only feature found to be important across all three algorithms. *Abbreviations: ADI_N-Area Deprivation Index (national percentile), PHQ-9-Patient Health Questionnaire-9, Readiness_Relapse-Readiness to change relapse score, SSWS-self-selected walking speed, 6MWT-6-Minute Walk Test, LR-Logistic regression, SVM-Support vector machine, RF-Random forest*.

Figure 3A displays the model performances for the home versus community threshold. All models demonstrated a moderate level of test-set accuracy after optimizing for class weights, where LR and SVM were fit with balanced class weights and RF was fit with none. Note that for the home versus community classification problem, we used balanced accuracy to measure model performance, meaning the scores represent how the model accurately identified individuals on average across the home and community classes. Overall, RF performed the worst, achieving an average balanced accuracy score of 68.1% (SD 6.5%, range 50.6 – 87.3%) with selected features and 67.9% (SD 5.3%, range 58.6% - 82.7%) when using all features. SVM followed, achieving an average balanced accuracy score of 77.3% (SD 5.4%, range 62.7% - 90.6%) with selected features and 73.1% (SD 5.7%, range 60.8% - 84.1%) when using all features. Finally, LR achieved the best overall balanced accuracy with an average score of 78.6% (SD 4.8%, range 68.7% - 90.1%) with selected features and 75.6% (SD 5.5%, range 63.3% - 87.6%) when using all features. The similarities in model performance accuracies when comparing a model with all features to the simplified model following regularization across all algorithms demonstrate that the regularization phase was effective in reducing the number of features while maintaining model performance accuracy.

**Fig 3.**
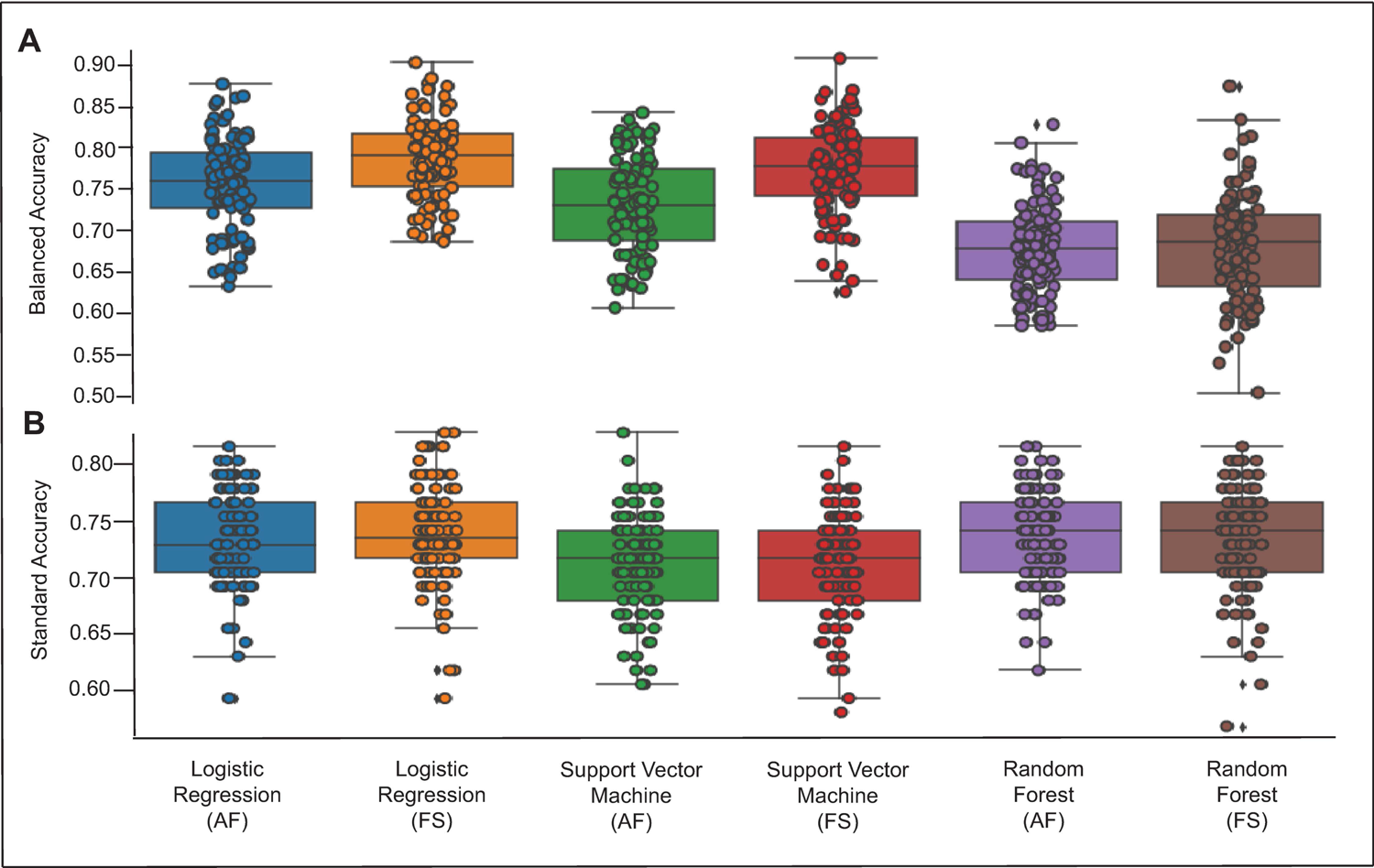
Model performance for the home vs. community threshold (2500 steps/day; A-upper figure) and aerobic threshold (5500 steps/day; B-bottom figure). Model performance for each algorithm is displayed with all features included (AF) and with feature selection (FS) that occurred as a result of the regularization step. Circles represent individual accuracy results for model performance during the 100 different train-tests splits of the data. Diamonds represent outliers. A higher accuracy score reflects better model performance.

### Results for aerobic threshold

After the regularization process, both linear SVM and LR produced the exact same results, with 16 features retained and carried forwards into the second stage of the analysis (see Y axis of Fig. 4).

**Fig 4.**
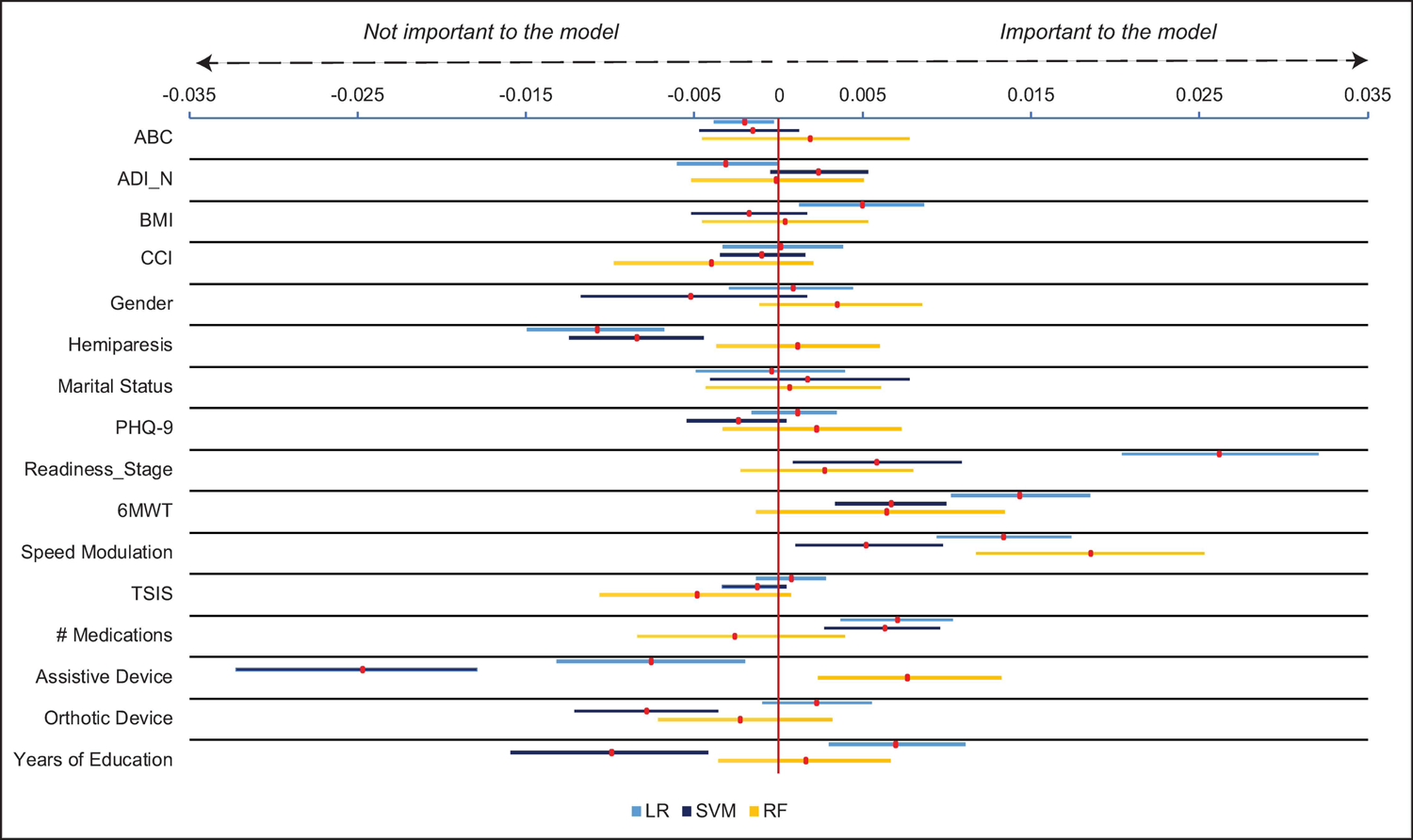
Drop column feature importance for aerobic threshold (5500 steps/day). Red markers show mean feature importance with 95% bootstrapped confidence interval. Speed modulation was the only feature found to be important across all three algorithms. *Abbreviations: ABC-Activities Specific Balance Confidence Scale, ADI_N-Area Deprivation Index (national percentile), BMI-body mass index, CCI-Charlson Comorbidity Index (age-adjusted), PHQ-9 (Patient Health Questionnaire-9), Readiness_Stage-Readiness to change stage score, 6MWT-6-Minute Walk Test, TSIS-time since initial stroke, LR-Logistic regression, SVM-Support vector machine, RF-Random forest*.

The drop column procedure was then run using these 16 variables over 100 random train-test splits of the data for all three algorithms. From these results, the only feature found to be important to all three algorithms was speed modulation, suggesting that the ability to change walking speed is critically important for predicting the aerobic step threshold in stroke. The full results of the drop column procedure for the aerobic threshold are displayed in Figure 4.

Figure 3B displays the model performances for the aerobic threshold. All models demonstrated a moderate level of test-set accuracy. Overall, SVM performed the worst, achieving an average standard accuracy score of 71.1% (SD 4.6%, range 58.0% - 81.5%) with selected features and 71.1% (SD 4.4%, range 60.5% - 82.7%) when using all features. LR and RF performed marginally better than SVM, with LR achieving standard accuracy scores of 73.9% (SD 4.5%, range 59.3% - 82.7%) with feature selection and 73.1% (SD 4.2%, range 59.3% - 81.5%) when using all features. RF achieved standard accuracy scores of 73.3% (SD 4.6%, range 6.8% - 81.5%) with feature selection and 73.5% (SD 4.1%, range 61.7% - 81.5%) when using all features. Again, the regularization phase was effective in reducing the number of features while maintaining model performance accuracy.

## Discussion

We determined that 6MWT was the only variable found to be an important predictor across all three algorithms in distinguishing home versus community ambulators. We also found that speed modulation was the only variable deemed important across all algorithms in distinguishing between stroke survivors who meet physical activity guidelines versus those who do not. Considering that 6MWT and speed modulation are measures of a stroke survivor’s physical capacity and were the only variables found to be important across all three mathematically unique algorithms led us to conclude that measures of physical capacity are *primary characteristics* that distinguish between groups of stroke survivors using these binary step thresholds.

Our finding that 6MWT was a primary characteristic for distinguishing between home versus community ambulators is in agreement with previous studies demonstrating that the 6MWT discriminates between functional walking categories in individuals with stroke [3, 26]. This finding is also aligned with several studies reporting that measures of physical capacity are, in general, important predictors of functional walking categories [3, 14, 26, 27] and physical activity [20] in individuals with stroke. As past work has generally utilized a single analytical approach, our results extend this work by demonstrating that the 6MWT prevails as an important predictor across multiple settings and further suggests that walking endurance is important for predicting whether a stroke survivor will achieve community mobility status using a 2500-step threshold.

These results are clinically important and suggest that clinicians should target the individual’s walking endurance to achieve the goal of community ambulation. However, as can be observed in Figure 2, additional features were deemed important across some algorithms, but not all. The Area Deprivation Index, Patient Health Questionnaire-9, readiness to change relapse score, self-selected walking speed, assistive device use, and years of education were found to be important for both SVM and LR, but not RF. Taken together, these findings suggest that addressing walking endurance is likely *necessary but not sufficient* for achieving a 2500-step threshold and that *ancillary features* (defined as predictors that were important to one or two algorithms, but not all three), including depressive symptoms and readiness to change activity behavior, may need to be addressed to fully achieve this step threshold.

For the aerobic threshold, speed modulation most consistently improved the prediction when using standard accuracy. Previous work has demonstrated that speed modulation is related to fall status [56] and daily walking activity [57] in older adults. Here we showed that speed modulation is also an important predictor of whether a stroke survivor will achieve a daily step threshold reflecting physical activity guidelines.

Since there is no uniform consensus on what magnitude of class imbalance warrants the use of standard versus balanced accuracy, we also assessed whether using balanced accuracy for the aerobic threshold would change our result. In this analysis, we found that both 6MWT and speed modulation were primary characteristics for achieving the aerobic step threshold (see S1 Fig). This similarity in result regardless of the use of standard accuracy (in which the 6MWT was close to being considered a primary characteristic) or balanced accuracy increases our confidence in our methodology and suggests that measures of physical capacity are critically important for achieving the aerobic step threshold.

However, our results demonstrate that targeting physical capacity is likely *necessary but not sufficient* for achieving a 5500-step threshold. This is supported by the fact that some algorithms, but not all, found that ancillary features were important in predicting the outcome. For example, both LR and SVM found that readiness to change activity behavior and the number of medications taken, were also important for predicting the aerobic step threshold, suggesting that readiness to change activity behavior and physical health status, may also need to be addressed to fully achieve this threshold.

We examined predictors of both 2500 and 5500-step thresholds as past work suggests that these thresholds likely provide different information. The 2500-step threshold is intended to distinguish between stroke survivors who walk primarily within their home setting from those who walk within their community [3]. As community ambulation typically involves greater walking distances [58], sufficient walking endurance is likely necessary to traverse community distances, lending credence to our finding that 6MWT is a primary characteristic of the home versus community threshold. In contrast, the 5500-step threshold is intended to distinguish between those who meet physical activity guidelines through walking and those who do not [10]. Physical activity guidelines are expressed in terms of exercise intensity and increasing one’s walking speed is one approach to increasing exercise intensity [9]. Therefore, it logically follows that the ability to modulate walking speed is a primary characteristic of those who meet intensity-based physical activity guidelines through walking. However, when using balanced accuracy for the aerobic threshold, we found the 6MWT was also a primary characteristic and increasing one’s exercise duration is another approach to meeting physical activity guidelines. We therefore conclude that measures of physical capacity are *primary characteristics* necessary to achieve these step thresholds and that associated *ancillary characteristics* may need to be addressed to fully achieve these step thresholds.

Importantly, these results should not be interpreted as the 6MWT and speed modulation represent the *best* single predictors of home versus community or aerobic step thresholds (see S2 Fig for predictive accuracy of 6MWT and speed modulation alone in predicting thresholds). This kind of inference would require determining that they hold the greatest predictive power over the other variables, which was not the purpose of this study. What our results do indicate, however, is that 6MWT and speed modulation are primary characteristics that distinguish stroke survivors who do and do not meet these step thresholds. Similarly, it is worth noting that neither 6MWT nor speed modulation had the greatest importance across all three algorithms in either classification setting (see Figs 2 and 4). Thus, our results extend upon literature examining predictors of stepping activity in stroke by demonstrating that there are features that are *primary characteristics that should be addressed* as well as *ancillary features that may need to be addressed* based on the unique circumstances of the individual.

With respect to model performance, RF was outperformed by SVM and LR in the home versus community threshold, with LR marginally achieving the best results over SVM. In the case of the aerobic threshold, RF and LR outperformed SVM with average accuracy scores within less than 1% of each other both with and without selected features. It is important to note that we did minimal hyperparameter tuning in this assessment, only optimizing the parameter for class weights in the case of the home versus community threshold due to the class imbalance. After tuning just this single hyperparameter, LR and SVM saw improvements in average performance of 7.3-13.5% after tuning, where RF’s performance did not improve with balanced class weights. It is possible that with more exhaustive hyperparameter tuning, model performance in the case of both thresholds could be improved.

Importantly, model performance accuracies were similar or better for a model with only features retained after regularization compared to a model with all features for all three algorithms and both thresholds. This validated the regularization phase, as we were able to reduce the number of variables to a relevant subset without compromising model performance. More directly, we were able to predict the home versus community and aerobic threshold classifications as well with 7 and 16 features, respectively, as we were when using all 25 features.

### Limitations

First, the class imbalance in the case of the home versus community threshold may have limited the performance of our algorithms. Consequently, model performance metrics across samples could suffer from high variability. In addition, with only 58 home ambulators, those in our sample may not be representative of the stroke population, as individuals were excluded if they walked at a self-selected gait speed of <0.3 m/s.

However, we aimed to address this limitation by using balanced accuracy as our metric of model performance for the home versus community threshold to avoid potential scenarios resulting in inflated measures of accuracy caused by the class imbalance (e.g., a constant model classifying all points as community ambulators). Second, although we used step thresholds endorsed by previous studies, we recognize that there are likely individuals who may not fit these exact criteria. For example, stroke survivors who achieve the step threshold to be considered community ambulators may take these steps primarily within their home environment.

## Conclusions/Implications

A stroke survivor’s physical capacity to walk is a *primary characteristic* that can be used to determine whether they will achieve step thresholds corresponding to home versus community ambulation and physical activity guidelines. However, measures of physical capacity were not necessarily the single best predictors of achieving these thresholds. Thus, addressing physical capacity is *necessary but not sufficient* for achieving these thresholds and *ancillary factors,* such as readiness to change activity behavior and physical health status, among others, may need to be addressed based on the individual’s unique clinical presentation. Future work on larger sample sizes that contain greater representation of home ambulators and other potentially relevant variables, such as fatigue and quality of life, is necessary.

## Data Availability

The data and code associated with this work are available on Open Science Framework at https://osf.io/tgzpb/

## Acknowledgements

None.

## Supporting information

## S1 Supplementary Material. Supplement_10-6-2021

### Machine learning algorithms

This section of the supplement is intended to provide a brief overview of the machine learning algorithms used in this study for readers unfamiliar with them. The three machine learning (ML) algorithms used in this analysis were Logistic Regression (LR), Support Vector Machine (SVM) with a radial basis function (RBF) kernel, and Random Forest (RF) classifier. These were chosen for this analysis because they are all commonly used supervised ML algorithms that are mathematically distinct from one another. A supervised classification algorithm is one that is trained by being provided with both data (X) and labels (Y), similar to a flash card. The way that supervised classification algorithms generally work is that they use the training data and labels to construct a function, f(x), to make predictions about which class other points in the same population belong to. Here, each point *x = (x^1^, x^2^, …, x^m^)* where *x^k^* is the value of *x* that corresponds with the k^th^ feature. This is done by solving an optimization problem to find the best parameters for f(x) based on what quantity is being optimized. The form of the predictor f(x) and the optimization process varies across ML algorithms.

In the case of LR with binomial class labels, as we have in our analysis, the general idea is that, based on our sample data, X, and labels, Y, we want to model the probability that Y=1, given X takes some value x, i.e.

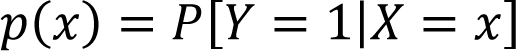

Then we also have that the probability that Y = 0 given X = x is 1-p(x). Now, to model this probability, p(x), LR uses the logistic function which takes values in (-∞, ∞) and maps them to the interval [0, 1] (the values of the probabilities). During this process, we want to find some set of parameters, β (the vector of beta weights), that fit the data in some optimal way where the logistic function to model this probability is:

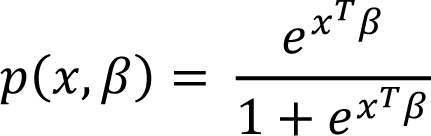

Given our training data has n observations, the goal of LR is find the values of β that maximize the log-likelihood function:

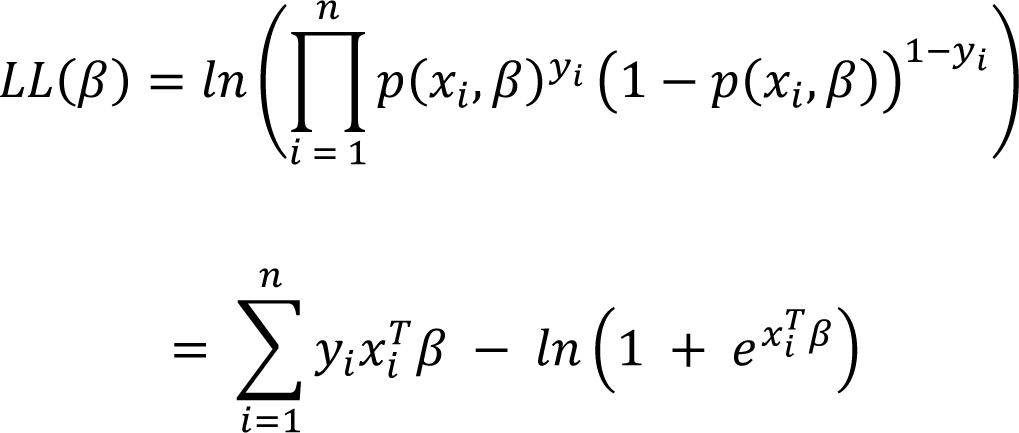

One important thing to note is that of the three models used in our analysis, LR is the most closely related to a linear model in the sense that the term *x^T^β* is a linear combination of the values of the vector x with the beta weights. This means that LR is only able to detect linear relationships between the features, where non-linear ones might exist.

For SVM, rather than computing the probability that a given observation belongs to some class, the goal is to establish boundaries with an allotted margin of error in the space the data comes from. These boundaries, which are defined by linear hyperplanes, separate the space the data comes from into regions. Then, a point would be classified depending on which of these regions the point lies in. In general, if our data set consists of m variables, then our data points live in an m-dimensional space. A hyperplane in an m-dimensional space would be defined by:

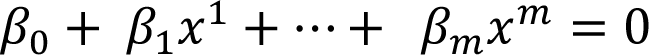

Which, in vector form, would be:

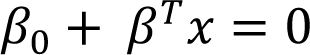

Where x is a single data point, β_0_ is the bias term, and *β* is the vector of beta weights. Notice that, similar to LR, the linear hyperplane in the m-dimensional space is only able to use linear relationships between the variables, but it is very possible that the data is not linearly separable. This is where one of the benefits of SVM, the use of a kernel, comes into play. A kernel, such as RBF, allows for additional “new” variables to be constructed using non-linear combinations of the existing variables. This allows the data to be projected into higher, m+d dimensional space, where d is the number of “new” variables, in which the data may now be separable by a linear hyperplane in this new, higher dimensional space. The way this would change the formulation of the boundary would be:

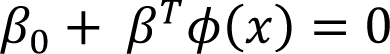

where Φ is an unknown function of *x* that is related to the transformation into higher dimensional space. The term Φ(x) is what would be the non-linear part of the definition of the hyperplane in *m*-dimentional space.

The RBF kernel, which is defined by:

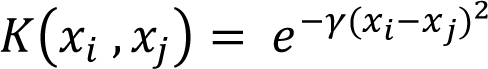

allows the data to be projected into an infinite dimensional space, but rather than ever actually making the physical transformation, the kernel functions allow for these higher dimensional relationships between the points to be computed because the kernel function acts as a dot product in that space. In this way, the RBF kernel can be thought of as a measure of how “close” two observations x_i_ and x_j_ are in infinite dimensional space. What makes SVM so robust is not only that it can use a kernel but also that, when establishing this boundary, there are allowances for misclassification of training data to handle noise. The optimization problem then becomes maximizing the distance between the points and the hyperplane while trying to minimize misclassification error:

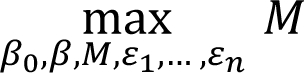

Subject to the constraints:

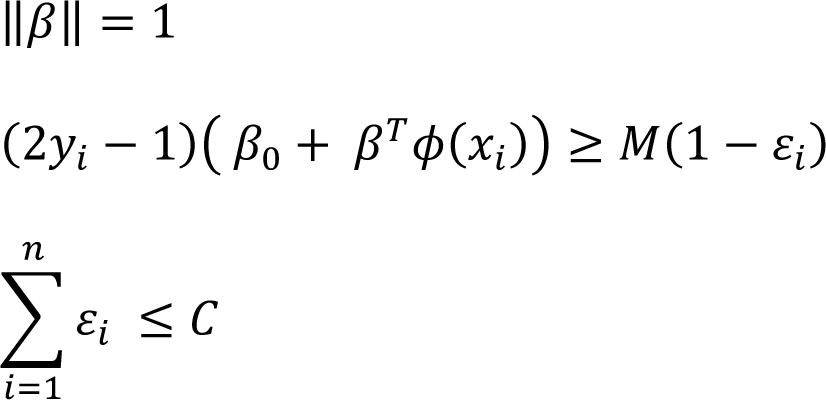

Where *M is the size of the margin, (2y_i_ — 1)(β_0_ + β^T^φ (x_i_))* is the distance from the observation *x_i_* to the hyperplane in the higher dimensional space, and C and ε_i_ ≥ 0 are the total error tolerance and the “slack” allowed for each observation, *x_i_*, respectively. This optimization problem can be solved using a Lagrange multiplier, which eliminates the dependency on the unknown function Φ(x). The end result is a potentially non-linear boundary that is as far away as possible from most points in each class, thus creating a robust classifier in the physical feature space.

Finally, RF is what is referred to as an “ensemble” because it is much more a collection of individual models, rather than a single model on its own. Simply put, a random forest is exactly what it sounds like: it is a collection (or “forest”) of decision trees which are each constructed using random subsets of the training data. Once a new point is passed into the RF model, each tree in the forest will classify that point and then RF will choose the class with the most “votes” in the forest. Each decision tree is made up of a collection of nodes that act like a path where each node is defined by a single feature with some threshold. The first node is the root node and connected to that are branches and then leaves. If we think of each of these nodes as possible stops on the tree’s path to a decision about a point, *x = (x^1^, x^2^, …, x^mm^)*, at each stop, the value of one of x’s entries will determine the next node it will stop at until it reaches a leaf node where the path ends and a decision is made based off of where it ended up. When growing each tree, a bootstrap sample of the training data is used and at each new node, a random subset of the variables is chosen, from which the best single predictor will be used to define the threshold that splits that node. This optimization of the individual decision tree is more of a greedy algorithm than an actual “optimization” problem in the classical sense. Consider an example where we are deciding how to best split a node with a random subset of k ≤ m features. For each feature *j* in {1, …, k} choosing a threshold, *t*, will create two regions, one for the points where the value of the feature j is less than or equal to *t* and one where the value is greater than *t*:

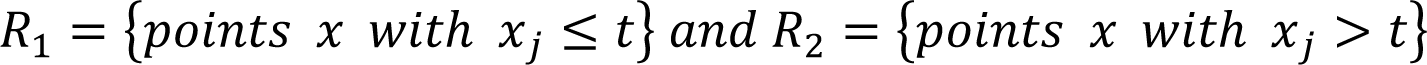

Then the feature j and the threshold *t* are chosen to be those that best classify the training data, i.e., those that minimize the classification error across the regions by satisfying:

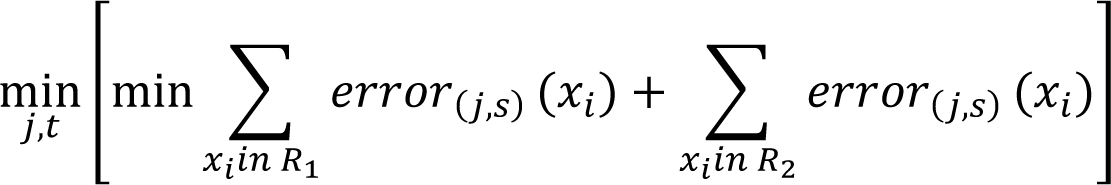

Where *error_R(j,s)_(x)* is the quantification of the error of classifying point x_ii_in region R using the variable j with threshold s. For classification, a popular metric to use here is the Gini Index. The end result is a robust model that is known to perform well in a wider variety of situations, but that is not very interpretable.

### Lasso regularization

In this analysis, Lasso (Least Absolute Shrinkage and Selection Operator) regularization was used to perform dimensionality reduction in the first phase of the feature importance process. This was done to reduce noise and redundancy among the full set of variables; thus, the goal was to reduce the number of variables while still retaining the same amount of information. As seen in the results section of this paper, this goal was achieved.

When fitting a linear model to data, as mentioned above, there is an optimization problem being solved. In general, for a linear model of the form:

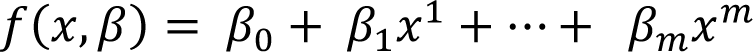

You want to find the values of the coefficients, β_j_, such that you are minimizing the loss function, *L(β)*. What Lasso regularization does is place a penalty on the size of the β coefficients where the optimization problem changes to finding the coefficients *β_1_, …, β_m_* such that we want to solve:

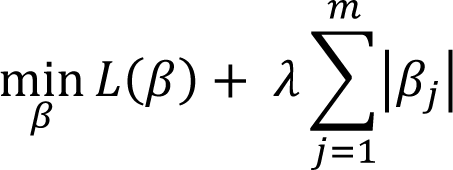

Where λ > 0 [1]. Note that in our analysis, the values of the regularization parameters can be interpreted as 1/λ.

The reason Lasso regularization was chosen for this analysis is that the penalty it places on the size of the β coefficients shrinks them in a way that forces some of the coefficients to 0 [1]. In doing this, the remaining features with non-zero coefficients can be interpreted as the subset of variables that were “chosen” by lasso regularization.

### Balanced accuracy for imbalanced data

We used two metrics of model performance throughout the analysis: standard accuracy in the case of the aerobic threshold (5500 steps/day), which is the total proportion of correctly classified points in the test set:

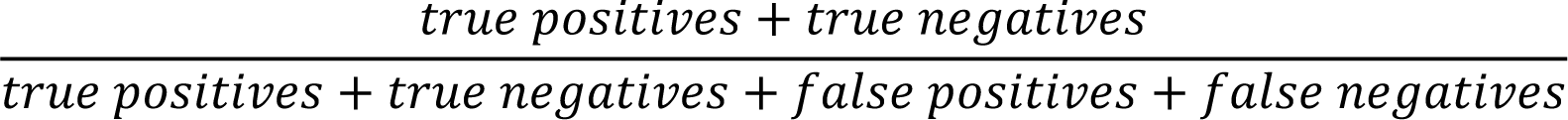

and balanced accuracy in the case of the home vs. community threshold (2500 steps/day), which is the arithmetic mean of the recall (or sensitivity) and specificity:

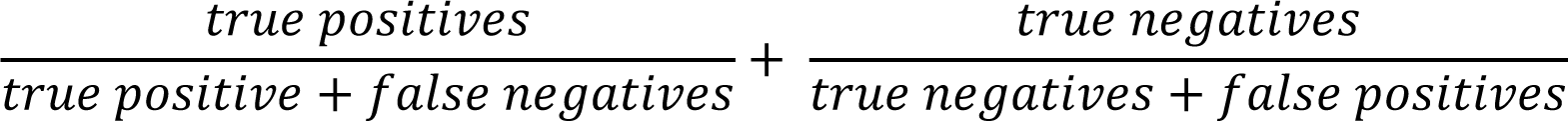

These metrics were chosen to ensure that the model performance metric used was representative of how well the model was performing on the target class.

The class distribution of each threshold was 58 (21.64%) home ambulators and 210 (78.36%) community ambulators in the case of the home vs. community threshold and 185 (69.03%) below the minimum aerobic activity threshold and 83 (30.97%) above for the aerobic threshold. While class distributions are not perfectly balanced in each case, the class imbalance in the case of the home vs. community threshold is starker at 1:4 and, more importantly, in favor of the negative class, unlike the aerobic threshold [2]. In the case of any significant class imbalance where the target class (the positive class) is the minority, the metric of standard accuracy could misrepresent how well the model is performing on the target class because standard accuracy may still be high, even if the model is not performing well on the target class [2]. In cases like this, we can use a metric like balanced accuracy, which allows the performance on the minority class to hold equal weight to that of the majority class and is also robust to misclassification noise [3].

### Results for the aerobic threshold (5500 steps/day) using balanced accuracy

When using the metric of balanced accuracy for the aerobic threshold, the optimized regularization parameters were 2.9 for LR and 0.99 for linear SVM. Given that when we use standard accuracy, the optimal regularization parameters were very similar at 2.9 and 0.9, it follows that the results of the lasso regularization stage when using balanced accuracy were identical to the 16 features reported in the primary analysis: 6MWT, speed modulation, CCI age-adjusted score, PHQ-9, readiness to change stage score, ABC, usual orthotic and assistive device, marital status, years of education, gender, BMI, side of hemiparesis, time since initial stroke, number of medications, and ADI_N.

In the second stage, the drop column procedure was run using these 16 features. For LR, 10 features were found to be important, 6 of which were the important features found when using standard accuracy. In order of importance, these features were: readiness to change stage score, speed modulation, 6MWT, usual assistive device, number of medications, usual orthotic device, years of education, BMI, CCI age-adjusted score, and time since initial stroke. For SVM, the same four features were found to be important as when using standard accuracy. In order of importance, these features were: readiness to change stage score, number of medications, speed modulation, and 6MWT. Like SVM, RF also had four features found to be important, which, in order of importance were: speed modulation, usual assistive device, 6MWT, and PHQ-9. From these results, speed modulation was still found to be a primary characteristic for the aerobic threshold, in addition to 6MWT. BMI, CCI, PHQ-9, readiness to change stage score, time since initial stroke, number of medications taken, assistive and orthotic device use, and years of education were found to be ancillary characteristics. Supplemental Figure 1 displays the results for the drop column procedure for the aerobic threshold using balanced accuracy.

Importantly, these results are very similar to the results found when using standard accuracy. Note that when standard accuracy was used, 6MWT was found to be important for both SVM and LR but the 95% confidence interval for RF was only barely negative, resulting in 6MWT not being found to be a primary characteristic. It is encouraging that these results were found in this analysis because it is reflective of the fact that, even with mild class imbalance, standard accuracy was an appropriate choice of metric for this threshold.

### 6-minute walk test as a single predictor of home vs. community thresholds (2500 steps/day)

A precision-recall curve was used to assess the predictive ability of the 6MWT alone in predicting the home vs. community threshold (2500 steps/day; Supplemental Figure 2A). Similar to an ROC analysis, a precision-recall curve can be used to assess the skill of a prediction model, where:

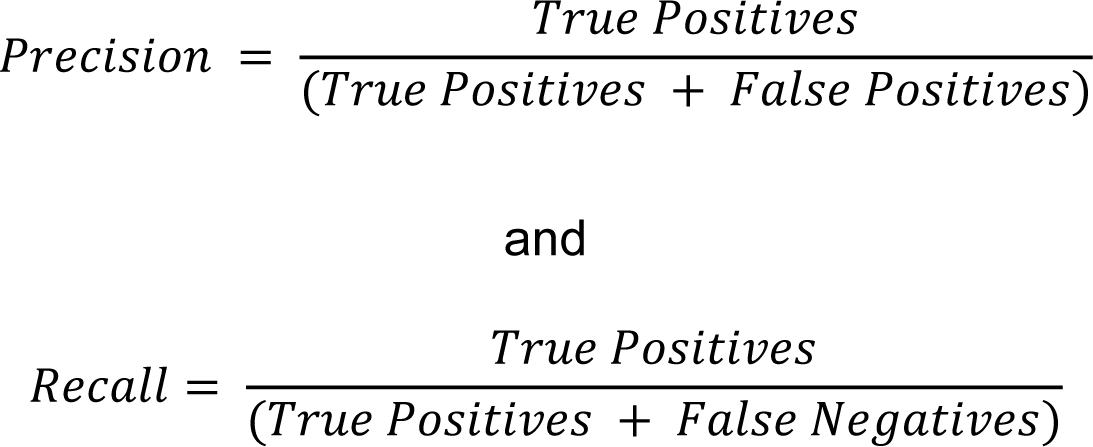

Recall is also referred to as sensitivity. As depicted in Supplemental Figure 2, a precision-recall curves plots the precision on the Y axis and recall on the X axis, where a point at (1,1) in the upper right corner of the figure would reflect a model with perfect precision and perfect recall. These curves can often be used instead of ROC curves in cases of class imbalance, as they are more sensitive to measures of recall. Similar to an ROC analysis, the area under the curve (AUC) can be computed to provide a measure of how well a model is performing where the AUC can have a maximum value of 1. A no-skill classifier (i.e., a model that randomly “guesses” the class) is depicted as a dashed line and changes based on the distribution of positive and negative cases:

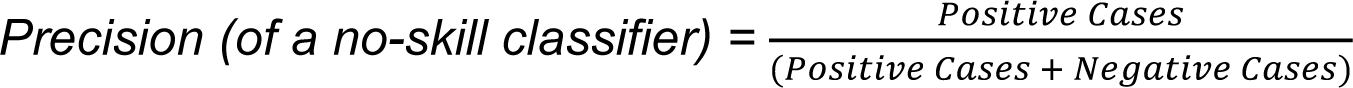

Previous work suggests that a precision-recall plot is more informative than an ROC plot when evaluating a binary classifier in the presence of imbalanced data [4, 5], as was the case with the home vs. community threshold. For comparison purposes, we present the precision-recall curves for the 6-Minute Walk Test and speed modulation for predicting home vs. community and aerobic thresholds, respectively (S2 Fig).

As shown in Supplemental Figure 2A, the AUC for the 6MWT in predicting home vs. community ambulation is 0.642 which exceeds that of a no-skill classifier (precision of no-skill classifier = 58/(210 + 58) = 0.2164).

### Speed modulation as a single predictor of aerobic threshold (5500 steps/day)

Supplemental Figure 2B displays the precision-recall curve for speed modulation in predicting the aerobic threshold of 5500 steps/day. The dashed line reflects a no-skill classifier and is calculated as: precision = 185/(83 + 185) = 0.6903. The AUC of 0.849 exceeds that of the no-skill classifier. When comparing the two precision-recall curves (and specifically the AUC values), it can be observed that speed modulation is a stronger predictor of the aerobic threshold than the 6MWT is in predicting the home vs. community threshold. We refer the reader to the reference list below for further reading on these topics.

---

**S1 Fig.**
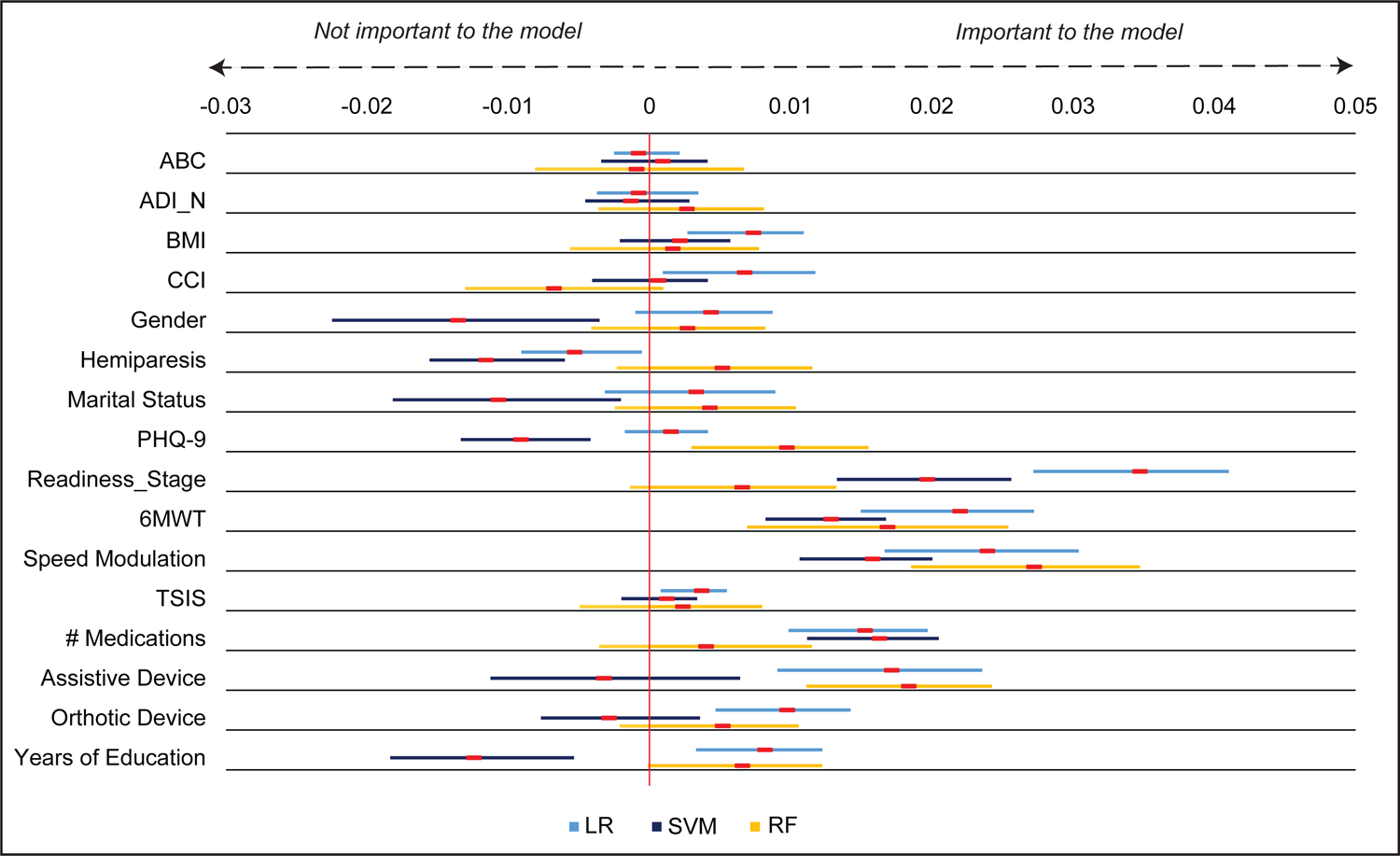
Drop column feature importance for aerobic threshold (5500 steps/day) using balanced accuracy. Red markers show mean feature importance with 95% bootstrapped confidence interval. 6MWT and speed modulation were the only features found to be important across all three algorithms. *Abbreviations: ABC-Activities Specific Balance Confidence Scale, ADI_N-Area Deprivation Index (national percentile), BMI-body mass index, CCI-Charlson Comorbidity Index (age-adjusted), PHQ-9-Patient Health Questionnaire-9, Readiness_Stage-Readiness to change stage score, 6MWT-6-Minute Walk Test, TSIS-time since initial stroke, LR-Logistic regression, SVM-Support vector machine, RF-Random forest*.

**S2 Fig.**
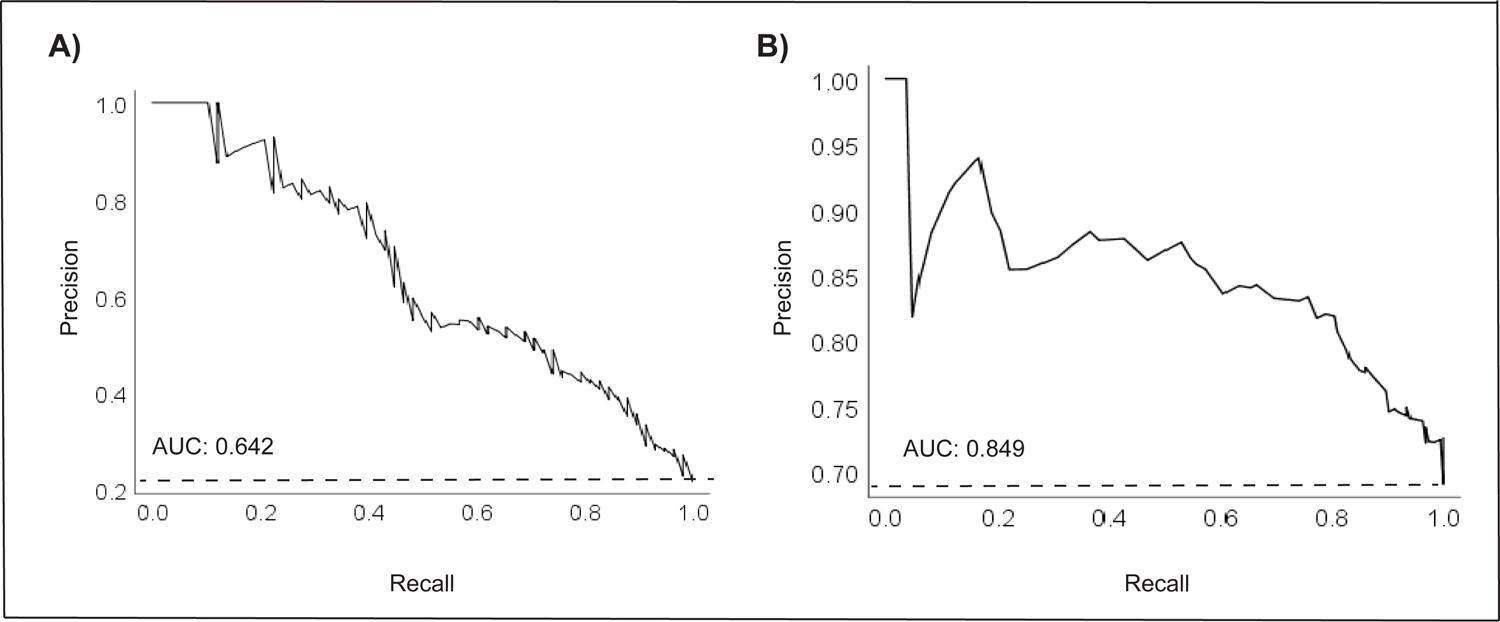
Precision-recall curve for predicting home versus community ambulation using the 6-minute walk test (A) and aerobic threshold using speed modulation (B). Precision is defined as: Precision = True Positives/(True Positives + False Positives). Recall is defined as: Recall = True Positives/(True Positives + False Negatives). The solid line plots the precision-recall curve, and the dash line reflects a no-skill classifier (a model that cannot discriminate between classes). *Abbreviations: AUC-Area Under Curve*.

